# Insulin resistance, and not β-cell impairment, mediates association between *Mycobacterium tuberculosis* sensitization and type II diabetes mellitus among US adults

**DOI:** 10.1101/2024.03.10.24304039

**Authors:** IM Magodoro, A Aluoch, B Claggett, MJ Nyirenda, MJ Siedner, KA Wilkinson, RJ Wilkinson, NAB Ntusi

## Abstract

Type 2 diabetes mellitus (T2DM) may be a long-term sequela of infection with *Mycobacterium tuberculosis (M.tb)* by mechanisms that remain to be fully explained. We evaluated association between *M.tb* sensitization and T2DM among U.S adults and, via formal mediation analysis, the extent to which this association is mediated by insulin resistance and/or β-cell failure. These evaluations accounted for demographic, socio-economic, behavioral and clinical characteristics. T2DM was assessed by fasting plasma glucose, 2-hour oral glucose tolerance testing and HbA1c; homoeostasis model assessment 2 (HOMA2) was used to estimate β-cell dysfunction (HOMA2-B) and insulin resistance (HOMA2-IR); while *M.tb* sensitization status was ascertained by tuberculin skin testing (TST). Exposure to *M.tb* was associated with increased risk for T2DM, likely driven by an increase in insulin resistance. Definitive prospective studies examining incident T2DM following tuberculosis are warranted.

**Research in Context:** *What is already known about this subject?:*

- Accumulating evidence suggests that pre-diabetes and new-onset type 2 diabetes mellitus (T2DM) may be a long-term complication of exposure to *Mycobacterium tuberculosis* (*M.tb*) via mechanisms that remain to be unraveled

*What is the key question?:*

- To what extent do insulin resistance and β-cell failure mediate the association between *M.tb* sensitization with T2DM among US adults?

*What are the new findings?:*

- *M.tb* sensitization is characterized by distinct glucose metabolic disturbances manifesting as increased risk of T2DM and isolated impaired fasting glucose (IFG)
- Insulin resistance, and not β-cell impairment, likely independently mediate the observed diabetogenic effects of *M.tb* sensitization

*How might this impact on clinical and/or public health practice in the foreseeable future?:*

- If corroborated by prospective studies, both TB programs and individual clinical care must incorporate monitoring of serum glucose and long-term metabolic outcomes
- This will be particularly urgent in sub-Saharan Africa and South-East Asia where scarce health resources coincide with overlapping endemic TB and epidemic T2DM

## Background

Tuberculosis (TB) is a long-acknowledged complication of type 2 diabetes mellitus (T2DM).^1^ In recent times, however, T2DM has come to the fore as possibly a sequel of both latent and active infection with *Mycobacterium tuberculosis* (*M.tb*).^2, 3, 4^ Heightened interest in the latter arises from the recognition that classical causes of T2DM, such as obesity, incompletely account for the diseasès high incidence. This has prompted the search for novel diabetes risk factors especially in low-and-middle income countries (LMICs)^5^ and among indigenous and minority communities in high-income settings (HICs).^6^ For example, diabetes in lean (body mass index (BMI) <25 kg/m^2^) individuals accounts for up to 32-60% of cases of T2DM in sub-Saharan Africa (SSA) and Southeast Asia (SEA).^5, 7, 8^ This contrasts sharply with HICs where at least 80% of T2DM patients are overweight/obese (BMI ≥25 kg/m^2^).^9^ With 120 million of the global 537 million prevalent T2DM cases (2021)^10^, SSA and SEA are also coincidentally the seat of endemic TB^11^. Despite the significance of TB as a likely T2DM risk factor, the pathophysiological mechanisms of this susceptibility remain unknown. These gaps imply that current clinical and public health strategies to prevent and control T2DM, or to mitigate the long-term consequences of tuberculosis, may be inadequate.

The two final pathophysiological pathways to T2DM development are islet β-cell failure and insulin resistance.^9, 12^ Their respective causes are multiple and overlap. Among others, they include disordered inflammatory responses, lipid metabolism and gut microbiome for insulin resistance^9^ and adverse early life exposures, oxidative stress, inflammatory cytokines and amyloid deposition for β-cell failure.^13^ Because T2DM is a heterogenous disease, the relative etiologic importance of β-cell failure and insulin resistance varies, as does their relative timing.^12^ , ^14^, The contributory roles of each of these two key pathways to new-onset T2DM associated with mycobacterial infection remain to be elucidated. TB pancreatitis, i.e., direct infection of pancreas tissue with *M.tb*, is uncommon while overt T2DM subsequent to it is very rare. However, *M.tb* infection likely has systemic-mediated diabetogenic effects.^16^ For example, the proinflammatory cytokine cascade set off by *M.tb* antigens may drive insulin resistance in skeletal muscle, adipose tissue and liver by adversely altering cellular and intracellular insulin signaling^16, 19, 20^, while islet amyloid deposition associated with *M.tb* infection may precipitate β-cell failure^18, 19^ through loss of both β-cell mass and function.^20^ Of note, amyloidosis in tuberculosis is documented in the USA^21^, and tuberculosis is the commonest cause of secondary amyloidosis in LMIC settings.^22^

Therefore, detailed mechanistic studies of TB’s diabetogenic potential remain an important and urgent priority. Here, we leveraged individual-level data from the 2011-2012 US National Health and Nutrition Examination Survey (NHANES) to address this challenge. Specifically, we assessed the association between *M.tb* sensitization and T2DM, and evaluated the extent to which insulin resistance and β-cell failure are key mechanisms through which *M.tb* infection leads to higher T2DM risk.

## Methods

We followed the guidelines of the Strengthening the Reporting of Observational Studies in Epidemiology (STROBE) in the conduct and reporting of our analyses.^23^

### Study design and participants

Details on methods, study protocols and ethical approvals of the NHANES are available elsewhere.^24^ Briefly, the NHANES is a recurring series of biennial cross-sectional surveys of the non-institutionalized US population. Participants are selected through multistage probability cluster sampling to ensure a nationally-representative sample. Individual-level data on health status and its determinants are collected through questionnaires, physical examination, and laboratory testing. All NHANES data are de-identified and publicly accessible, obviating the need for institutional review board approvals for any data analysis. For the present study, we used data from the 2011–2012 NHANES cycle for the main analysis, and the 1999-2000 NHANES cycle for the sensitivity analysis as explained below. All adults aged at least 20 years with complete data on fasting plasma glucose and insulin, oral glucose tolerance testing (OGTT) and tuberculin skin testing (TST) were eligible for inclusion in both analyses. Excluded were those with self-reported administration of insulin and/or extreme values of fasting/prandial glucose (<3 mmol/L or >25 mmol/L) or insulin (<20 pmol/L or >300 pmol/L).

### Study covariates, exposures, and outcomes

#### Covariates

Data on participants’ age, sex, race/ethnicity, socio-economic status, alcohol use, blood pressure (BP), BMI, waist circumference, serum cotinine and past medical diagnoses including auto-immune conditions (asthma, psoriasis, celiac disease, arthritis and thyroiditis) were extracted. Race/ethnicity was self-reported as Hispanic, non-Hispanic white, non-Hispanic black, and non-Hispanic Asian/Other. The family income-to-poverty ratio (PIR) was used to assess the socio-economic status with PIR ≤1.3 considered the poverty threshold [Sebelius 2011]. We defined tobacco exposure as “none” if serum cotinine <10 ng/mL, “passive exposure or light smoker” if serum cotinine ≥10 ng/mL and <300 ng/mL, and “heavy smoker” if serum cotinine ≥300 ng/mL.^25^ Participants who did not have at least 12 alcohol-based drinks in the preceding year or ever, and those who had at least 12 alcohol-based drinks in their lifetime but not in the past year were classified as “non-drinkers”. On the other hand, participants who had at least 12 drinks in the past year were defined as current drinkers, and were further classified as “heavy current drinkers” if they reported ever having 4/5 or more drinks every day, or “light/moderate current drinkers” if not.^26^ Lastly, we extracted data on blood pressure, BMI, and waist circumference measurements.

#### M.tb sensitization status

*M.tb* sensitization was ascertained by tuberculin skin testing (TST) using tuberculin-purified protein derivative (PPD) product, Tubersol^®^ (Sanofi, Bridgewater, NJ). Skin induration was measured 48-72 hours after placement of PPD. Similar TST testing and quality control methods were followed in the two NHANES cycles.^24^ Because neither chest radiographs nor tuberculosis symptoms screening were completed in the NHANES, skin induration ≥10mm was considered indicative of *M.tb* sensitization.^27^ Data on BCG vaccination status were also not available.

#### Pancreatic islet β-cell function and insulin resistance

Homoeostasis model assessment (HOMA) 2 estimates of β-cell function (HOMA2-B), and insulin resistance (HOMA2-IR) were calculated using fasting plasma glucose and insulin with the HOMA calculator (University of Oxford, Oxford, UK).^28^

#### Diabetes mellitus and prediabetes states

The ADA criteria (2023) were used to define glycemic status.^29^ Diabetes mellitus was defined as any of fasting plasma glucose (FPG) ≥7.0 mmol/L, 2-hour OGTT plasma glucose (prandial plasma glucose) ≥11.1 mmol/L or glycated hemoglobin (HbA1c) ≥6.5%. Data were not available to distinguish types 1 and 2 diabetes mellitus. However, the incidence of type 1 diabetes, which is insulin dependent, peaks in puberty.^30^ Menke *et al.,* (2013) using 1999-2000 NHANES data estimated type 1 diabetes to be 4.8% of all diabetes in the US.^31^ Because we included participants aged ≥20 years old and not currently using insulin we therefore presumed all identified diabetes cases in our study to be T2DM. We tested the robustness of this assumption in sensitivity analyses that used a higher age cut-off (≥40 years old) when the prevalence of undiagnosed type 1 diabetes is likely to be low. Among non-diabetics, we also defined prediabetes as any of HbA1c ≥5.6% and <6.5% or fasting plasma glucose ≥5.6 mmol/L and <7 mmol/L or prandial plasma glucose ≥7.8 mmol/L and <11.1 mmol/L. We similarly defined, among non-diabetics, isolated impaired fasting glucose (isolated IFG) as fasting plasma glucose between 5.6 and 7.0 mmol/L and PPG <7.8 mmol/L; and isolated impaired glucose tolerance (isolated IGT)was fasting plasma glucose <7.0 mmol/l and prandial plasma glucose 7.8 and 11.1 mmol/L.

### Data analysis

Analyses were conducted using R, version 3.6.3 (R Foundation for Statistical Computing, Vienna, Austria) and Stata version 17.0 (StataCorp, College Station, TX, USA). All probability values were 2- sided with p-values <0.05 considered indicative of statistical significance. We did not apply sampling weights to our analyses. First, socio-demographic, behavioral, and clinical characteristics of the cohort were summarized by *M.tb* sensitization status. The distribution of participants’ glucose metabolism indices were then plotted by *M.tb* sensitization status, and their differences assessed by quantile regression models. The β-coefficients (95% CI) from these models were reported as mean difference in the median value. Next, probit regression models with postestimation margins were applied to determine the association between *M.tb* sensitization and diabetes and prediabetes states and results presented graphically. Models were adjusted for age, sex, race/ethnicity, family poverty-income ratio, alcohol consumption, tobacco exposure, waist circumference, and self-reported auto-immunity.

The last step was mediation analysis using the counterfactual framework^32^ to examine whether, and how much, insulin resistance or β-cell failure contributed to the association of *M.tb* sensitization with T2DM (**Supplementary Figure 1).** We used the “*mediation*” package in R^33^ and analyses were performed by including one mediator at a time. Confounding factors for the mediation effect were the same as those included in the regression analyses. We modeled (50^th^ percentile) insulin resistance or β-cell function using quantile regression (mediator model) and (2) T2DM using probit regression (outcome model). Interactions between mediators and exposure (*M.tb* sensitization/HOMA2-B, and *M.tb* sensitization/HOMA2-IR) were tested, and if statistically significant, were included in the mediation analyses.

#### Sensitivity analysis

The sensitivity analyses entailed two steps. First, we repeated the causal mediation analysis using the 2011-2012 NHANES cycle but including only participants aged ≥40 years old. The second step, however, used the 1999-2000 NHANES cycle with similarly defined inclusion criteria (age ≥20 years old; not using insulin) and variables as the primary analysis.

## Results

### Characteristics of study participants

The analytic sample included 1,843 adults (≥20 years old) with complete exposure (*M.tb* status) and outcomes data (T2DM, HOMA indices). Missing covariates data were less than 2%, and thus we proceeded with complete case analysis. Their baseline characteristics are summarized in **Table 1**.

**Table 1.**
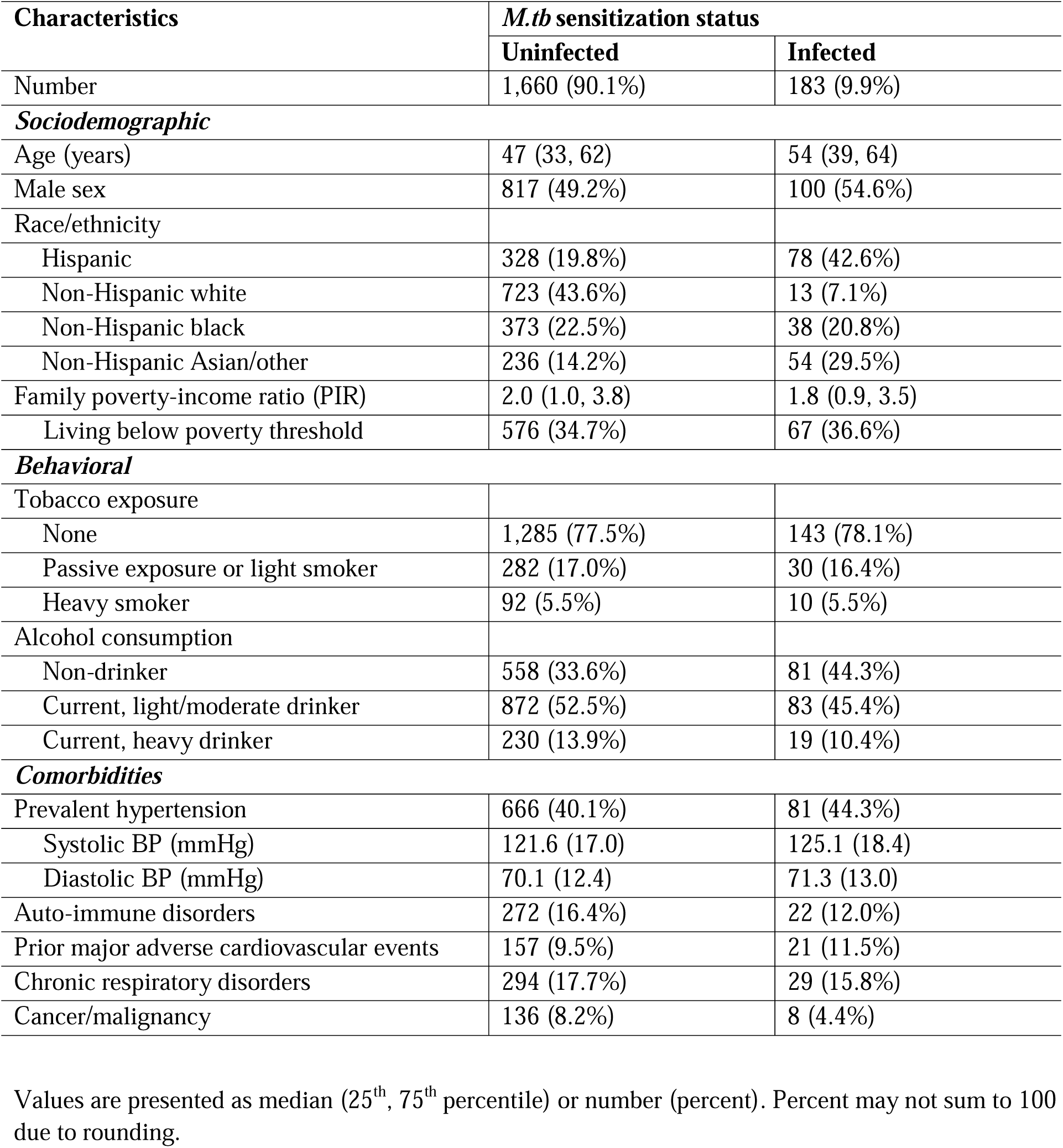
Participants’ characteristics according to *M.tb* sensitization status, unweighted US NHANES 2011-2012 sample.

| Characteristics | *M.tb* sensitization status |  |
| --- | --- | --- |
|  | Uninfected | Infected |
| Number | 1,660 (90.1%) | 183 (9.9%) |
| ***Sociodemographic*** |  |  |
| Age (years) | 47 (33, 62) | 54 (39, 64) |
| Male sex | 817 (49.2%) | 100 (54.6%) |
| Race/ethnicity |  |  |
| Hispanic | 328 (19.8%) | 78 (42.6%) |
| Non-Hispanic white | 723 (43.6%) | 13 (7.1%) |
| Non-Hispanic black | 373 (22.5%) | 38 (20.8%) |
| Non-Hispanic Asian/other | 236 (14.2%) | 54 (29.5%) |
| Family poverty-income ratio (PIR) | 2.0 (1.0, 3.8) | 1.8 (0.9, 3.5) |
| Living below poverty threshold | 576 (34.7%) | 67 (36.6%) |
| ***Behavioral*** |  |  |
| Tobacco exposure |  |  |
| None | 1,285 (77.5%) | 143 (78.1%) |
| Passive exposure or light smoker | 282 (17.0%) | 30 (16.4%) |
| Heavy smoker | 92 (5.5%) | 10 (5.5%) |
| Alcohol consumption |  |  |
| Non-drinker | 558 (33.6%) | 81 (44.3%) |
| Current, light/moderate drinker | 872 (52.5%) | 83 (45.4%) |
| Current, heavy drinker | 230 (13.9%) | 19 (10.4%) |
| ***Comorbidities*** |  |  |
| Prevalent hypertension | 666 (40.1%) | 81 (44.3%) |
| Systolic BP (mmHg) | 121.6 (17.0) | 125.1 (18.4) |
| Diastolic BP (mmHg) | 70.1 (12.4) | 71.3 (13.0) |
| Auto-immune disorders | 272 (16.4%) | 22 (12.0%) |
| Prior major adverse cardiovascular events | 157 (9.5%) | 21 (11.5%) |
| Chronic respiratory disorders | 294 (17.7%) | 29 (15.8%) |
| Cancer/malignancy | 136 (8.2%) | 8 (4.4%) |

Values are presented as median (25<sup>th</sup>, 75<sup>th</sup> percentile) or number (percent). Percent may not sum to 100 due to rounding.

Compared to individuals without *M.tb*, those with *M.tb* sensitization were older [median (25^th^, 75^th^ percentile): 54 (39, 64) vs. 47 (33, 62) years old] and more frequently Hispanic (42.6% vs. 19.8%) and male (54.6% vs. 49.2%). Rates of family poverty (PIR <1.3: 36.6% vs. 34.7%), tobacco exposure and alcohol use were comparable between the two groups. There were also no notable differences in rates of self-reported prevalent comorbidities although participants with *M.tb* sensitization had higher systolic BP (125.1 vs. 21.6 mmHg) than their counterparts without.

Glucose metabolism profiles of participants are summarized in **Figure 1a-f**. *M.tb* sensitization was associated with worse profiles of fasting [median (25^th^, 75^th^ percentile): 5.6 (5.2, 6.3) vs. 5.4 (5.1, 6.0) mmol/L; p=0.007] (F**igure 1d**) and prandial plasma glucose [6.6 (5.2, 8.8) vs. 5.9 (4.8, 7.3) mmol/L; p=0.048) (**Figure 1e**), although the latter marginally so. Similarly, HbA1c was higher with *M.tb sensitization* [5.7 (5.3, 6.2)%] than without [5.5 (5.2, 5.9)%; p=0.004] (**Figure 1f**).

**Figure 1.**
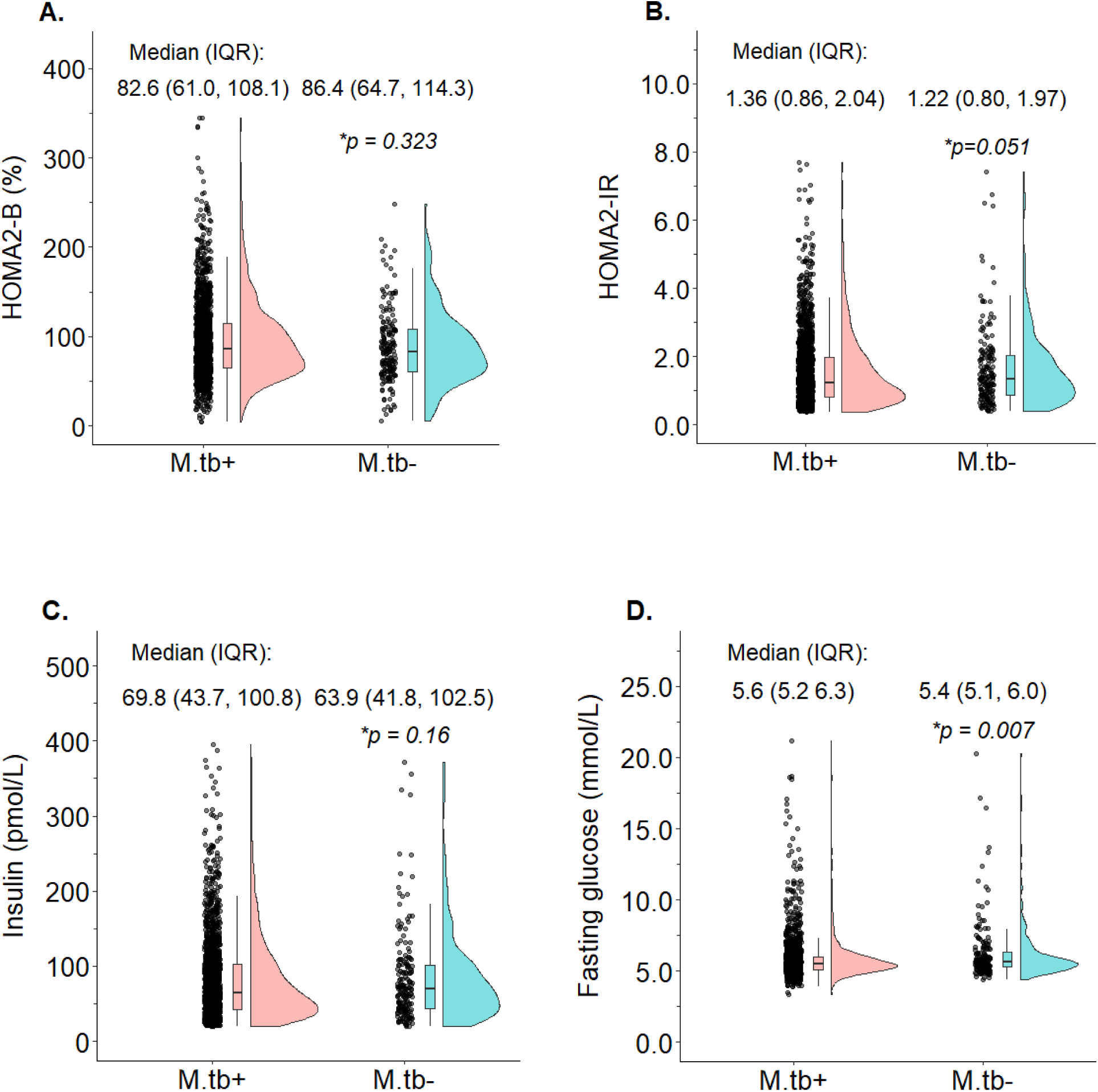
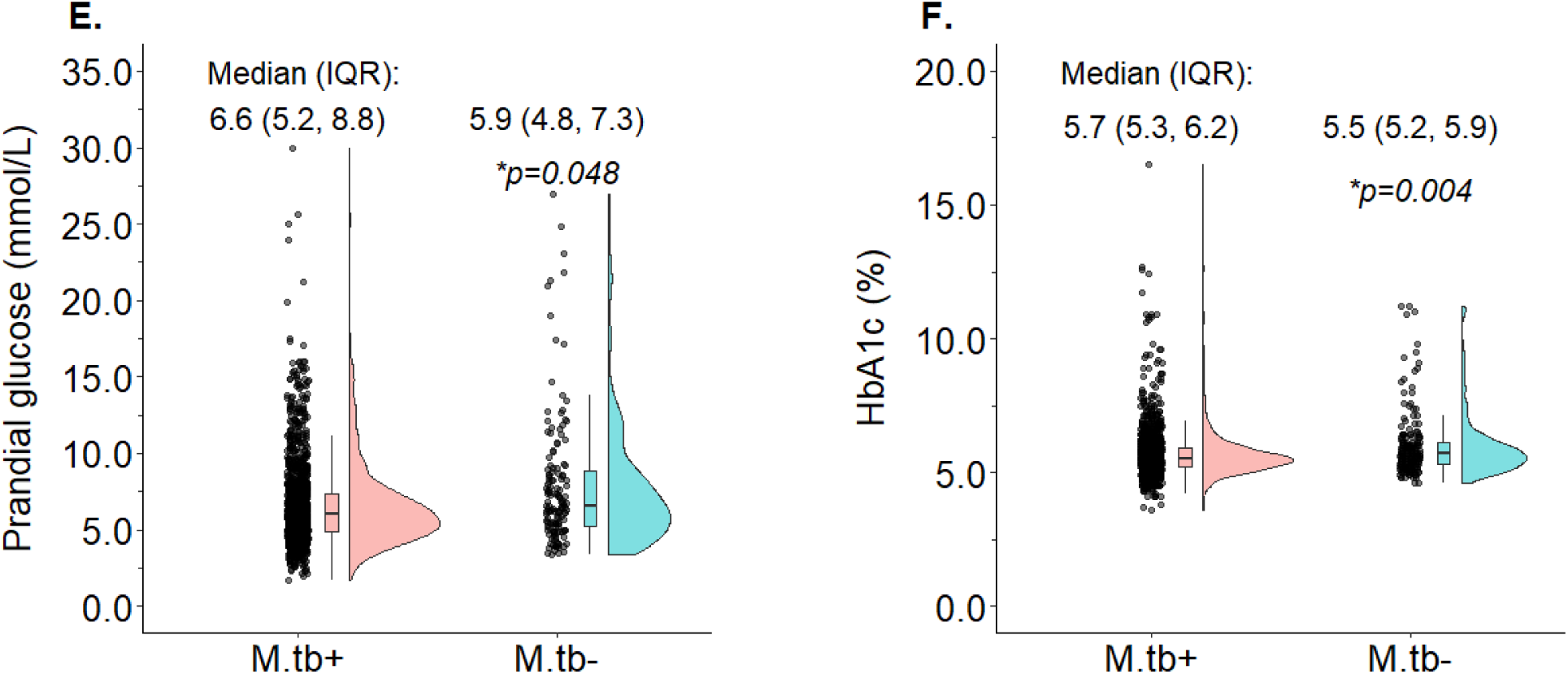
Participants’ cardiometabolic characteristics according to *M.tb* sensitization status, unweighted US NHANES 2011-2012 sample. *M.tb*+ and *M.tb*- represent, respectively, *M.tb* sensitized and *M.tb* uninfected. * P-values derived from (unadjusted) quintile regression models of difference in median (50^th^ percentile) value.

### Distribution of pancreatic islet β-cell function, insulin and insulin resistance

The distribution of β-cell function and insulin resistance is summarized in **Figures 1a-b** and **Tables 2** and **3**, and **Supplementary Table 1**. The association between insulin resistance and *M.tb* sensitization was statistically significant (**Table 2**) with and without confounder adjustment (**Figure 1b** and **Table 2**). For example, the adjusted mean difference [AMD (95% CI)] in median HOMA2-IR between the two groups was 0.16 (0.03, 0.29) (p=0.014), being higher for those with *M.tb* sensitization. The differences in β-cell function [AMD in median: -3.1 (-10.5, 4.3); p=0.42] and fasting plasma insulin [AMD in median : 1.66 (- 5.00, 8.33); p=0.63] (**Supplementary Table 1** and **Figure 1c**) were negligible as these results did not reach statistical significance. The other significant determinants of increased insulin resistance were female (vs. male) sex, older age, greater central adiposity, and alcohol abstention (**Table 2**). In contrast, male (vs. female) sex, older age and living in poverty were significantly correlated with impaired β-cell impairment (**Table 3**).

**Table 2.**
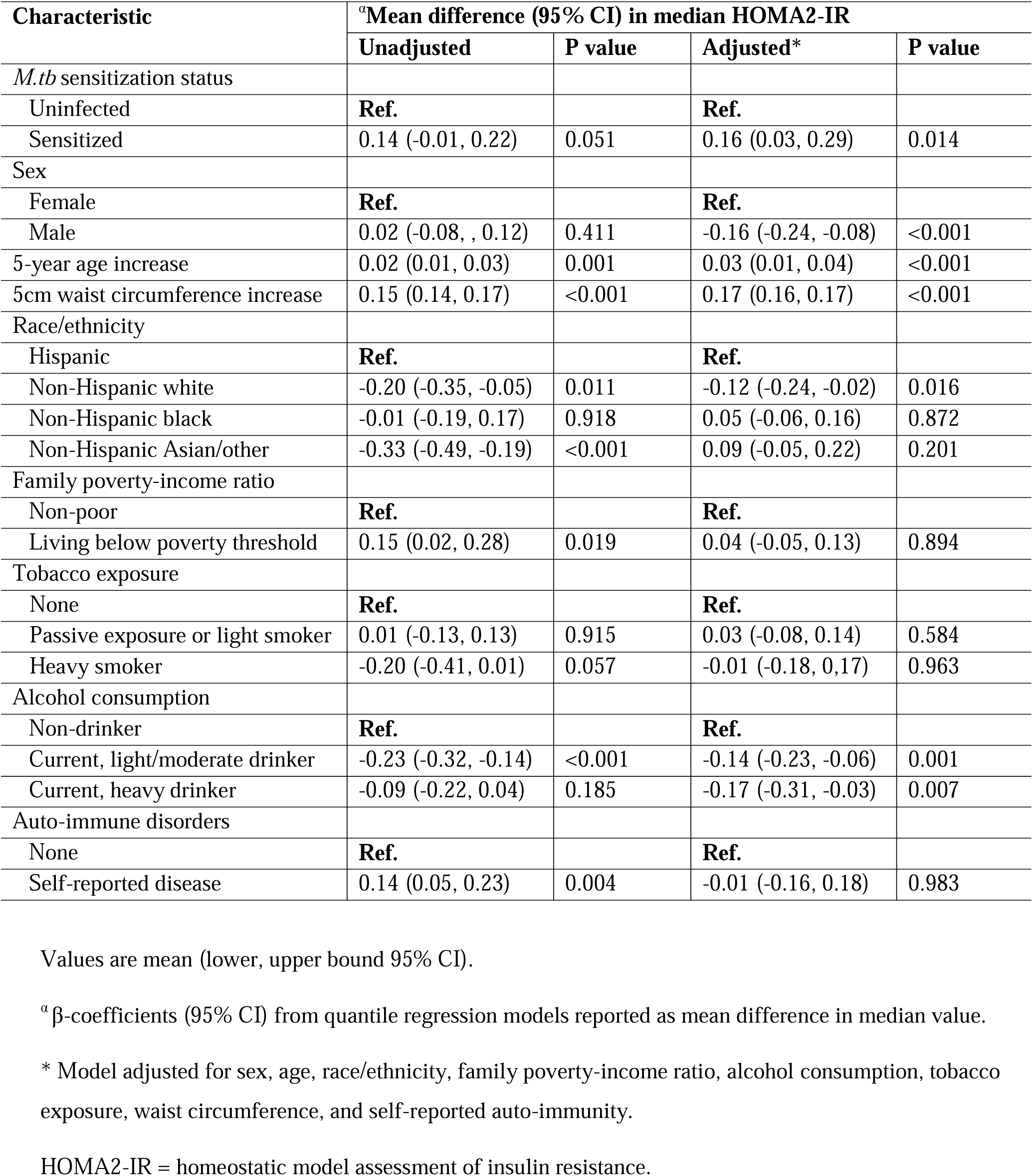
Associations between *M.tb* sensitization and insulin resistance, unweighted US NHANES 2011-2012 sample.

**Table 3.** Associations between *M.tb* sensitization and pancreatic β-cell function, unweighted US NHANES 2011-2012 sample.

| Characteristic | <sup>a</sup>Mean difference (95% CI) in median HOMA2-B |  |  |  |
| --- | --- | --- | --- | --- |
|  | Unadjusted | P value | Adjusted* | P value |
| *M.tb* sensitization status |  |  |  |  |
| Uninfected | **Ref.** |  | **Ref.** |  |
| Sensitized | -3.8 (-9.6, 2.0) | 0.201 | -3.1 (-10.5, 4.3) | 0.421 |
| Sex |  |  |  |  |
| Female | **Ref.** |  | **Ref.** |  |
| Male | -9.90 (-14.3, -5.51) | <0.001 | -11.3 (-14.5, -8.02) | <0.001 |
| 5-year age increase | -2.43 (-3.17, -1.68) | <0.001 | -3.22 (-3.01, -2.73) | <0.001 |
| 5cm waist circumference increase | 4.51 (3.87, 5.15) | <0.001 | 5.44 (4.91, 5.97) | <0.001 |
| Race/ethnicity |  |  |  |  |
| Hispanic | **Ref.** |  | **Ref.** |  |
| Non-Hispanic white | -2.30 (-7.46, 2.86) | 0.381 | 0.68 (-3.31, 4.66) | 0.762 |
| Non-Hispanic black | 3.31 (-2.94, 9.54) | 0.302 | 4.56 (-1.40, 10.5) | 0.134 |
| Non-Hispanic Asian/other | -8.72 (-14.1, -3.22) | 0.002 | 1.31 (-3.19, 5.81) | 0.571 |
| Family poverty-income ratio |  |  |  |  |
| Non-poor | **Ref.** |  | **Ref.** |  |
| Living below poverty threshold | 2.10 (-2.67, 6.87) | 0.384 | -3.73 (-6.97, -0.50) | 0.024 |
| Tobacco exposure |  |  |  |  |
| None | **Ref.** |  | **Ref.** |  |
| Passive exposure or light smoker | -1.20 (-7.28, 4.89) | 0.687 | -1.68 (-6.97, 3.26) | 0.510 |
| Heavy smoker | -7.80 (-18.6, 2.96) | 0.690 | -5.46 (-12.0, 1.06) | 0.101 |
| Alcohol consumption |  |  |  |  |
| Non-drinker | **Ref.** |  | **Ref.** |  |
| Current, light/moderate drinker | -3.70 (-8.64, 1.24) | 0.151 | -3.61 (-8.01, 0.80) | 0.391 |
| Current, heavy drinker | -4.40 (-12.6, 3.83) | 0.298 | -3.18 (-10.5, 4.08) | 0.393 |
| Auto-immune disorders |  |  |  |  |
| None | **Ref.** |  | **Ref.** |  |
| Self-reported disease | 1.70 (-4.32, 7.72) | 0.583 | 0.36 (-4.36, 5.08) | 0.512 |

Values are mean (lower, upper bound 95% CI).
<sup>a</sup> β-coefficients (95% CI) from quintile regression models reported as mean change in median value.
\* Model adjusted for sex, age, race/ethnicity, family poverty-income ratio, alcohol consumption, tobacco exposure, waist circumference, and self-reported auto-immunity.
HOMA2-B = homeostatic model assessment of β-cell function.

### Prevalence of diabetes and prediabetes states

T2DM was significantly more prevalent with *M.tb* sensitization compared to being *M.tb* uninfected in unadjusted (28.4% vs. 16.6%; p=0.002) (**Figure 2a**) and adjusted (25.7% vs. 16.7%; p<0.001) (**Figure 2b**) comparisons. Among the prediabetic states, only isolated IFG differed significantly by *M.tb* status. It was more prevalent among the *M.tb* sensitized [adjusted prevalence (95%CI): 21.4 (19.4, 23.6)%] than the uninfected [14.3 (9.2, 19.4)%; p=0.008] (**Figure 2b**).

**Figure 2.**
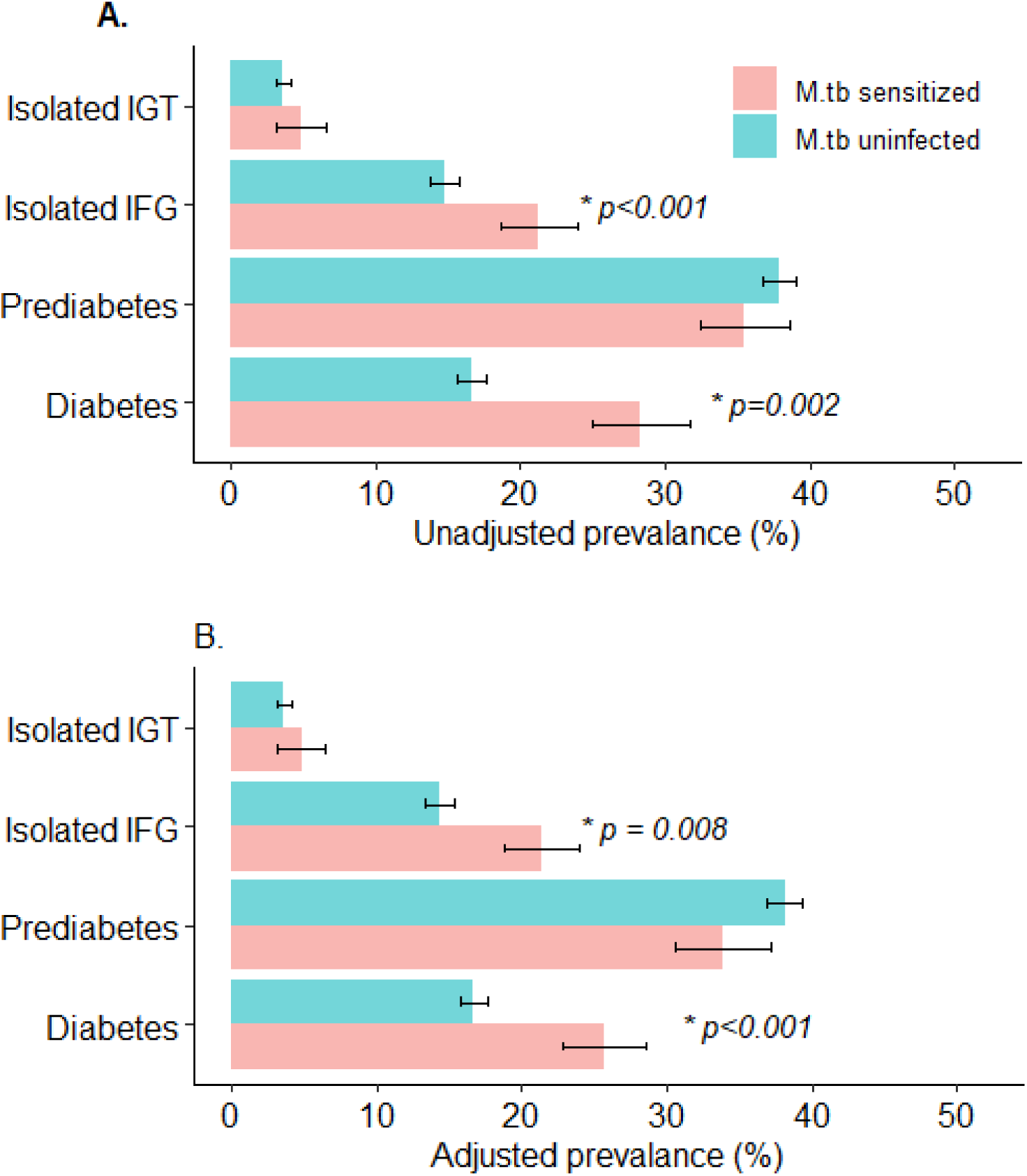
(a) Unadjusted and (b) adjusted prevalence of diabetes mellitus and prediabetic states according to *M.tb* sensitization status, unweighted US NHANES 2011-2012 sample. * P values are for *M.tb* sensitized versus *M.tb* uninfected comparisons, and are shown only if statistically significant (i.e., p<0.05) Prevalence adjusted for sex, age, race/ethnicity, family poverty-income ratio, alcohol consumption, tobacco exposure, waist circumference, and self-reported auto-immunity. Prediabetes = defined among non-diabetics as any of HbA1c ≥5.6% and <6.5% or fasting plasma glucose ≥5.6 mmol/L and <7 mmol/L or prandial plasma glucose ≥7.8 mmol/L and <11.1 mmol/L. Isolated IFG = isolated impaired fasting glucose; Isolated IGT = isolated impaired glucose tolerance; LTBI = latent tuberculosis infection.

### Mediated effects of islet β-cell function and insulin resistance

Exposure-mediator interactions were not statistically significant (*M.tb* sensitization/HOMA2-B: p=0.43; *M.tb* sensitization/HOMA2-IR: p=0.97) were not statistically significant, and thus were not included in the mediation models. Compared to being uninfected, *M.tb* sensitization was associated with greater risk of prevalent T2DM [adjusted absolute risk difference (adjusted ARD) (95%CI): 9.34 (2.38, 15.0); p<0.001] (**Table 4**). Of this total effect, 18.3 (3.29, 36.0)% - corresponding to an adjusted ARD of 1.65 (0.31, 3.00)% - was due to mediation by insulin resistance. In contrast, the pathway via β-cell function was not significant [adjusted ARD: 0.57 (-0.87, 2.0)%; p=0.48], and thus β-cell function did not meaningfully contribute to association between *M.tb* sensitization and T2DM [proportion mediated: 6.33 (-10.8, 21.0)%; p=0.50]. The magnitude and direction of these findings were replicated when the mediation analysis was repeated using either the 1999-2000 NHANES cycle or the 2011-2012 NHANES cycle but with higher inclusion age (≥40 vs. ≥20 years old) (**Table 4**).

## Discussion

Data from the present study demonstrate that *M.tb* sensitization is characterized by distinct glucose metabolic disturbances independent of age, sex and other potential determinants. We found evidence associating *M.tb* sensitization with increased risk of T2DM and isolated impaired fasting glucose (IFG) in US adults. In tandem, we also observed fasting and prandial hyperglycemia, as well as elevated HbA1c with *M.tb* sensitization. Insulin resistance, and not β-cell impairment, mediated the observed diabetogenic effects of *M.tb* sensitization. Because our study was cross-sectional, we make these inferences with caution. Definitive prospective mechanistic studies of incident T2DM following *M.tb* exposure are required. Notwithstanding, our findings do suggest that sharpened focus on long-term metabolic outcomes of tuberculosis patients is warranted. This is particularly urgent in SSA and SEA where scarce health resources must meet increasingly overlapping endemic TB and epidemic T2DM.

Exposure to *M.tb* represents an immuno-pathological spectrum.^34^ At one end is asymptomatic immune sensitization to mycobacterial antigens as evidenced by reactive TST and/or interferon (IFN)-gamma release assays (IGRAs) in apparently healthy persons. Clinically overt, and oftentimes fatal, TB disease lies at the opposite pole. How T2DM risk varies along this spectrum is unknown. Whereas we investigated impaired glucose regulation in *M.tb* sensitization, most studies to date have focused on active TB and report glucose intolerance in up to half (16.5-49%) of the cases.^35^ The features of impaired glucose regulation associated with *M.tb* sensitization in our study were fasting and (marginally) prandial hyperglycemia, and in turn isolated IFG. According to the literature, the majority of patients with glucose intolerance during active TB regress to normoglycemia after antituberculosis treatment.^16^ Transient hyperglycemia is also seen with other severe systemic infections, such as community-acquired pneumonia, supporting the possibility of a non-specific stress response to infection.^17^ Unfortunately, our study lacked follow-up data on the trajectories of *M.*tb-associated impaired glucose regulation. It is important to note, however, that these preclinical metabolic states independently predict future new-onset T2DM in the general population.^9^

A drawback of most available studies to date is the fact that their cross-sectional design is also compatible with undetected T2DM being present prior to the onset of TB. Recognizing this bi-directionality of the TB-T2DM relationship^2^, we nevertheless proceeded from the hypothesis that *M.tb* sensitization was diabetogenic. Our mediation analyses followed the counterfactual framework which - subject to necessary assumptions of positivity, consistency, and no unmeasured confounding for the exposure-outcome, mediator-outcome, and exposure-mediator relationships - provides valid causal estimates.^32^The total effect of exposure on outcome is decomposed into direct and indirect effects through a mediator. The direct effect captures the effect of exposure on outcome if the path via mediators is prevented or removed hypothetically. Our finding that *M.tb* sensitization was a risk factor for T2DM is thus consistent with the available albeit limited prospective studies of new-onset T2DM following *M.*tb exposure. Pearson *et al.,* (2018)^4^ reported a fivefold higher T2DM incidence among individuals with a history of clinical TB using UK-wide primary care data, while Magee *et al.,* (2022)^36^ found that US veterans with reactive TST and/or IGRA had up to 1.3 times higher risk of new-onset T2DM than their non-reactive peers. On the other hand, Young *et al.,* (2010) found no evidence of increased post-tuberculosis T2DM risk in a prospective cohort in Oxford, England.^37^

From a mechanistic standpoint, studies in both humans and animals provide evidence for the diabetogenicity of *M.tb*.^16^ For example, in the murine model of tuberculosis, *M.tb* has been shown to cause insulin resistance via dysregulation of lipid metabolism with ectopic deposition of fat in the liver and skeletal muscles.^38^ Insulin resistance has also been attributed to impaired liver function due to the toxic effect of anti-tuberculosis drugs.^16^ Inflammation from *M.tb* infection results in an environment of sustained pro-inflammatory cytokine production often leading to metabolic dysregulation and eventually insulin resistance.^16^ Philips *et al*., 2017 in South Africa found insulin resistance in a quarter of newly diagnosed male and female TB patients.^39^ Similarly, we found *M.tb* sensitization to be associated with greater insulin resistance, which in turn was on the mechanistic pathway to T2DM. In contrast, we did not find evidence for a role for β-cell impairment. The putative link between exposure to *M.tb* and β-cell impairment will be pancreatic amyloid deposition leading to loss of islet mass and function.^18, 19^ In fact, *M.tb* sensitized participants in our study had lower HOMA2-B than their uninfected counterparts although the differences did not reach statistical significance. This could also be attributed, at least in part, to our exclusion of participants on insulin, some of whom could have significant insulin deficiency.

### Strengths and limitations

Our study is among the first to explore the mechanistic roles of islet β-cell failure and insulin resistance in the diabetogenicity of *M.tb*, and thus sheds novel insights into a challenge of growing clinical and public health concern. The NHANES samples are drawn to reflect the diversity of the US population. Compared to currently available studies that are mostly health facility-based, our results therefore have greater generalizability. Data on TB-related symptoms, chest radiographs and sputum examinations in conjunction with TST would have enabled better stratification of the *M.tb* sensitized into those who have eliminated TB infection, controlled TB infection and subclinical TB infection.^34^ The cross-sectional design of our study remains a limitation and warrants caution with inferences as do likely residual confounding and potential misclassification biases. TST, for example, can register false positive results due to sensitization by environmental mycobacteria and/or BCG. Unfortunately, we did not have BCG vaccination data to adjust the TST cut-off value. Neither did we have data to robustly distinguish type 1 diabetes from T2DM. However, the consistency of our primary and sensitivity analyses does give some reassurance.

## Conclusion

Definitive prospective studies examining incident T2DM following *M.tb* exposure are urgently required, especially in SSA and SEA. Future studies should aim for more granular definition of *M.tb* sensitization using imaging, laboratory testing and clinical examination, and more accurate definitions of T2DM. Notwithstanding, our findings suggest that exposure to *M.tb* may be a novel risk factor for T2DM, likely driven by an increase in insulin resistance.

## Data Availability

All data in the present analysis are available at https://www.cdc.gov/nchs/nhanes/about_nhanes.htm

## Acknowledgements

RJW and KAW are funded by the Francis Crick Institute which is supported by Cancer Research UK (FC2112), Medical Research Council (FC2112) and Wellcome (FC2112). RJW also receives support from Wellcome (203135). For the purposes of open access the authors have applied a CC-BY public copyright to any author-accepted manuscript arising from this submission. MJS receives support from the US National Institutes of Health (K24 HL166024). NABN gratefully acknowledges support from the South African Medical Research Council, National Research Foundation, the US National Institutes of Health, Medical Research Council (UK), and the Lily and Ernst Hausmann Trust.

---

**Supplementary Table 1.** Associations between *M.tb* sensitization and insulin, unweighted US NHANES 2011-2012 sample.

| Characteristic | <sup>a</sup>Mean difference (95% CI) in median fasting plasma insulin |  |  |  |
| --- | --- | --- | --- | --- |
|  | Unadjusted | P value | Adjusted* | P value |
| *M.tb* sensitization status |  |  |  |  |
| Uninfected | Ref. |  | Ref. |  |
| Sensitized | 5.82 (-1.30, 12.9) | 0.112 | 1.66 (-5.00, 8.33) | 0.693 |
| Sex |  |  |  |  |
| Female | Ref. |  | Ref. |  |
| Male | -1.32 (-5.92, 3.28) | 0.571 | -7.98 (-11.84, -4.12) | <0.001 |
| 5-year age increase | 0.81 (0.20, 1.42) | 0.009 | -1.43 (-1.89, -0.96) | <0.001 |
| 5cm waist circumference increase | 7.65 (7.03, 8.28) | <0.001 | 8.63 (7.96, 9.30) | <0.001 |
| Race/ethnicity |  |  |  |  |
| Hispanic | Ref. |  | Ref. |  |
| Non-Hispanic white | -10.0 (-17.2, -2.89) | 0.006 | -5.85 (-10.94, -0.77) | 0.024 |
| Non-Hispanic black | 0.24 (-8.40, 8.88) | 0.906 | 3.96 (-2.08, 9.99) | 0.109 |
| Non-Hispanic Asian/other | -16.5 (-23.9, -9.12) | <0.001 | 4.63 (-1.43, 10.6) | 0.135 |
| Family poverty-income ratio |  |  |  |  |
| Non-poor | Ref. |  | Ref. |  |
| Living below poverty threshold | 6.66 (0.56, 12.8) | 0.032 | 1.27 (-2.96, 5.50) | 0.546 |
| Tobacco exposure |  |  |  |  |
| None | Ref. |  | Ref. |  |
| Passive exposure or light smoker | 0.66 (-5.99, 7.31) | 0.842 | 1.03 (-4.36, 6.42) | 0.721 |
| Heavy smoker | -9.36 (-20.3, 1.67) | 0.096 | 0.62 (-5.10, 6.33) | 0.833 |
| Alcohol consumption |  |  |  |  |
| Non-drinker | Ref. |  | Ref. |  |
| Current, light/moderate drinker | -11.6 (-16.2, -7.13) | <0.001 | -6.71 (-11.1, -2.36) | 0.003 |
| Current, heavy drinker | -5.76 (-12.2, 0.65) | 0.078 | -8.98 (-16.2, -1.76) | 0.015 |
| Auto-immune disorders |  |  |  |  |
| None | Ref. |  | Ref. |  |
| Self-reported disease | 7.14 (1.87, 12.4) | <0.001 | 1.21 (-5.51, 7.92) | 0.744 |

Values are mean (lower, upper bound 95% CI).
<sup>a</sup> β-coefficients (95% CI) from quantile regression models reported as mean difference in median value.
\* Model adjusted for sex, age, race/ethnicity, family poverty-income ratio, alcohol consumption, tobacco exposure, waist circumference, and self-reported auto-immunity.
HOMA2-IR = homeostatic model assessment of insulin resistance.

**Supplementary Figure 1.**
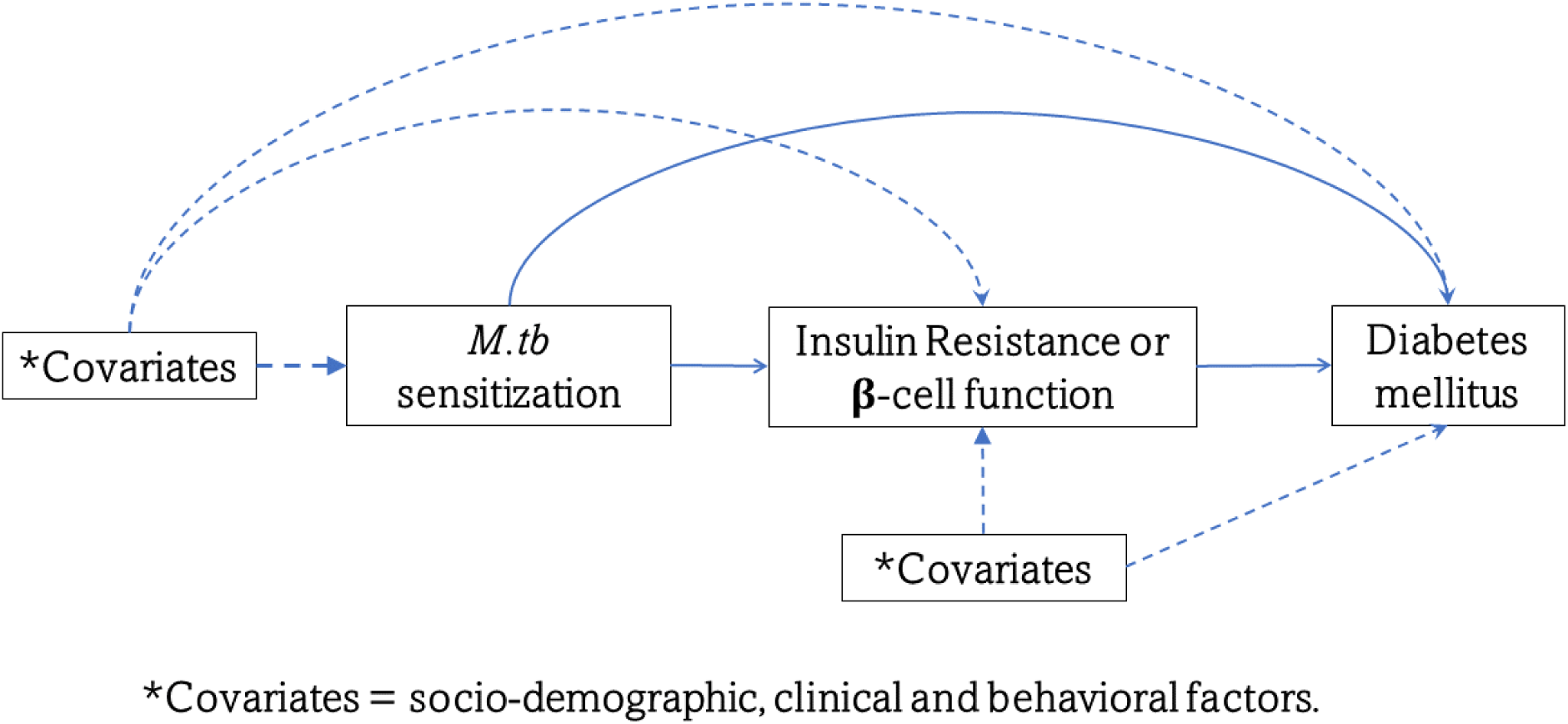
Directed acyclic graph (DAG) of the association to *M.tb* sensitization n and diabetes mellitus.

