## Supplementary material for "Insulin resistance, and not β-cell impairment, mediates association between *Mycobacterium tuberculosis* sensitization and type II diabetes mellitus among US adults": Table 4

**Table 4. Mediated effects of insulin resistance and β-cell function in the association between *M.tb* sensitization and diabetes mellitus, unweighted US NHANES 2011-2012 sample.**

| **Mediator** | **Adjusted*^,α^ absolute risk difference, % (95%CI)** | | | | | | **Proportion mediated** | |
| --- | --- | --- | --- | --- | --- | --- | --- | --- |
|  | **Total effect** | **P value** | **Direct effect** | **P value** | **Indirect effect** | **P value** | **% (95%CI)** | **P value** |
| ***HOMA2-IR*** |  |  |  |  |  |  |  |  |
| *Primary analysis* |  |  |  |  |  |  |  |  |
| NHANES 2011-2012;  ≥20 years old | 9.34 (2.38, 15.0) | <0.001 | 7.68 (1.43, 12.0) | <0.001 | 1.65 (0.31, 3.00) | 0.020 | 18.3 (3.29, 36.0) | 0.020 |
| *Sensitivity analysis* |  |  |  |  |  |  |  |  |
| NHANES 2011-2012;  ≥40 years old | 10.8 (7.42, 22.0) | <0.001 | 9.31 (4.86, 19.0) | <0.001 | 1.62 (1.45, 3.00) | <0.001 | 13.5 (11.4, 38.0) | <0.001 |
| NHANES 1999-2000 | 10.7 (3.5, 18.6) |  | 8.9 (2.0, 16.1) |  | 1.8 (0.4, 3.4) |  | 16.7 (9.4, 48.7) |  |
| ***HOMA2-B*** |  |  |  |  |  |  |  |  |
| *Primary analysis* |  |  |  |  |  |  |  |  |
| NHANES 2011-2012;  ≥20 years old | 9.04 (4.60, 16.0) | <0.001 | 8.47 (4.58, 14.0) | <0.001 | 0.57 (-0.87, 2.00) | 0.481 | 6.33 (-10.8, 21.0) | 0.501 |
| *Sensitivity analysis* |  |  |  |  |  |  |  |  |
| NHANES 2011-2012;  ≥40 years old | 11.6 (4.67, 19.0) | <0.001 | 11.5 (4.30, 19.0) | <0.001 | 0.0 (-1.4, 3.00) | 0.562 | 0.89 (-0.96, 22.0) | 0.560 |
| NHANES 1999-2000 | 12.5 (4.6, 20.6) |  | 11.9 (4.9, 19.4) |  | 0.5 (-2.1, 3.3) |  | 4.5 (2.8, 12.3) |  |

* Adjusted for sex, age, race/ethnicity, family poverty-income ratio, alcohol consumption, tobacco exposure, waist circumference, and self-reported auto-immunity.

^α^ Absolute risk difference between *M.tb* sensitized and *M.tb* uninfected as reference group.

Primary analysis based on 2011-2012 NHANES cycle with inclusion age ≥20 years old.

Sensitivity analyses based on (i) 2011-2012 NHANES cycle with inclusion age ≥40 years old, and (ii) 1999-2000 NHANES cycle with inclusion age ≥20 years old.
